# Deep learning for predicting COVID-19 malignant progression

**DOI:** 10.1101/2020.03.20.20037325

**Authors:** Cong Fang, Song Bai, Qianlan Chen, Yu Zhou, Liming Xia, Lixin Qin, Shi Gong, Xudong Xie, Chunhua Zhou, Dandan Tu, Changzheng Zhang, Xiaowu Liu, Weiwei Chen, Xiang Bai, Philip H.S. Torr

## Abstract

As COVID-19 is highly infectious, many patients can simultaneously flood into hospitals for diagnosis and treatment, which has greatly challenged public medical systems. Treatment priority is often determined by the symptom severity based on first assessment. However, clinical observation suggests that some patients with mild symptoms may quickly deteriorate. Hence, it is crucial to identify patient early deterioration to optimize treatment strategy. To this end, we develop an early-warning system with deep learning techniques to predict COVID-19 malignant progression. Our method leverages clinical data and CT scans of outpatients and achieves an AUC of 0.920 in the single-center study and an average AUC of 0.874 in the multicenter study. Moreover, our model automatically identifies crucial indicators that contribute to the malignant progression, including Troponin, Brain natriuretic peptide, White cell count, Aspartate aminotransferase, Creatinine, and Hypersensitive C-reactive protein.

---

Since 2020, COVID-19 has had a fundamental effect on people’s lives. As of August 12, 2020, the number of COVID-19 infections in the world has soared to 20.2 million (20,162,474) with a mortality of 3.7% (737,417/20,162,474)^1^, which greatly challenges public medical systems. France and The United Kingdom have the highest mortality in the world, which is 15.8% (30,227/191,265) and 14.9% (46,526/312,793), respectively. In comparison, the mortality in some other countries is much lower, such as 4.2% (9,207/218,519) in Germany^1^.

One of the most important causes for such a difference in mortalities is early identification and active intervention of patients with mild symptoms to prevent deterioration^2^. Clinical observations^3,4^ suggest that although around 80% of COVID-19 patients are mild or asymptomatic, some of them may rapidly deteriorate. More importantly, studies^5^ have shown that over 60% of patients died once they progressed into a severe/critical stage. Thus this group of patients requires special attention and treatment in advance. The focus of this study therefore, as illustrated in Supplementary Figure 1, is on an accurate prediction of COVID-19 malignant progression, which is conducive to the timely intervention of clinicians, rational optimization of medical resources, and the effective operation of the entire medical system.

Current research mainly focuses on exploiting clinical variables ascertained at hospital admission or quantitative CT parameters for progression prediction, via using nomogram^6,7^, Light Gradient Boosting Machine (LightGBM)^8^, or LASSO regression^9^. However, the performance of those methods is still far from that required for practical use, for three reasons: 1) a manual quantization of feature patterns is required, which leads to information loss before analysis; 2) temporal cues are more or less ignored, but crucial to an accurate prediction; 3) chest CT scans and clinical data capture different characteristics of patients, but the complementarity between them is not fully leveraged.

To address above issues, we resort to AI techniques to deliver an accurate model for the prediction of COVID-19 malignant progression. Our model, based on deep learning methods^10-12^, effectively mines the complementary information in the static clinical data and the dynamic sequence of chest CT scans. It operates on raw data in an end to end manner, which means any manual design of feature patterns or interference of clinicians is not required. Moreover, our model automatically identifies crucial indicators that contribute to the malignant progression, including Troponin, Brain natriuretic peptide, White cell count, Aspartate aminotransferase, Creatinine, and Hypersensitive C-reactive protein.

In summary, our work presents an early warning system and targets early identification of COVID-19 malignant progression for reducing the patient stratification uncertainty, optimizing the diagnosis and treatment, increasing the efficiency of medical resource allocation, improving the emergency response capacity of the medical system, and ultimately decreasing the mortality. Comprehensive experiments on three cohorts demonstrate that our system using both clinical data and CT scans not only achieves the best performance in the internal validation (AUC: 0.920, 95% CI: [0.861, 0.979], cohort one), but more importantly, has robust generalization power in the external validations (AUC: 0.885, 95% CI: [0.847, 0.923], cohort two; AUC: 0.862, 95% CI: [0.789, 0.935], cohort three).

## Results

### Dataset statistics

All data enrolled in this retrospective study is obtained from two hospitals in Wuhan, including Wuhan Pulmonary Hospital and three branches of Tongji Hospital. The collection and use of these data from two hospitals were approved by their IRBs, respectively. 1,040 patients with mild COVID-19 pneumonia at admission are considered in our study, including 491 males and 549 females, aged 18 to 95 (57.51 ± 14.75). 32.3% of patients (336/1,040) malignantly progressed to a severe/critical stage during the hospitalization, while the remaining 67.7% (704/1,040) did not (Fig. 1). The selected data is divided into three cohorts, of which the cohort one is used for the single-center study, and the cohort two and the cohort three are used for the multicenter study. The clinical data in three cohorts is summarized in Table 1.

**Fig. 1.**
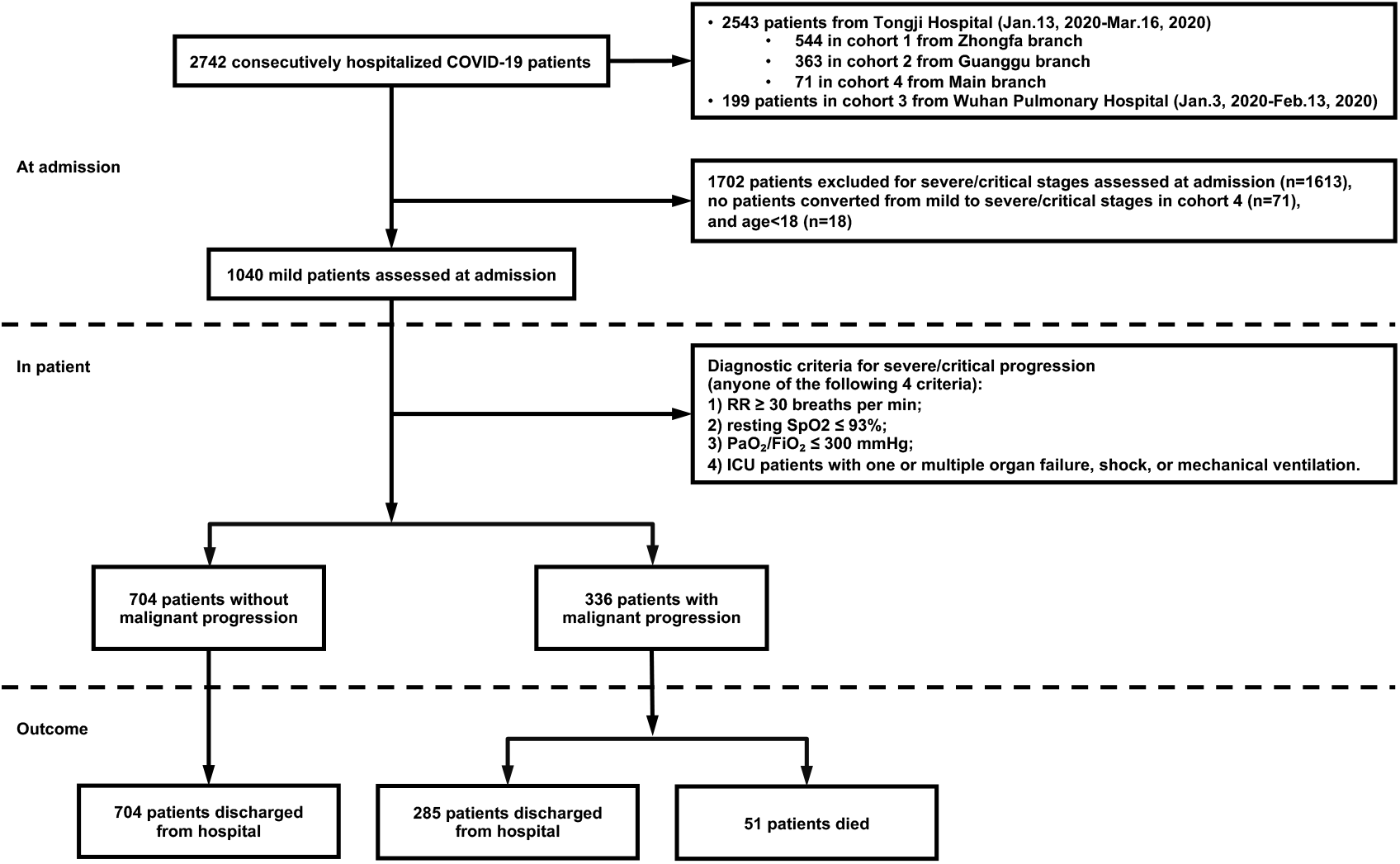
Flowchart of patient selection. A total of 1,040 out of 2,742 patients are selected according to the inclusion criteria. All the 1,040 patients have the complete clinical data required for the study and 57.9% of them underwent serial chest CT imaging. RR: respiratory rate, SpO_2_: blood oxygen saturation, PaO_2_: arterial oxygen partial pressure, FiO_2_: fraction of inspiration oxygen.

**Table 1.**
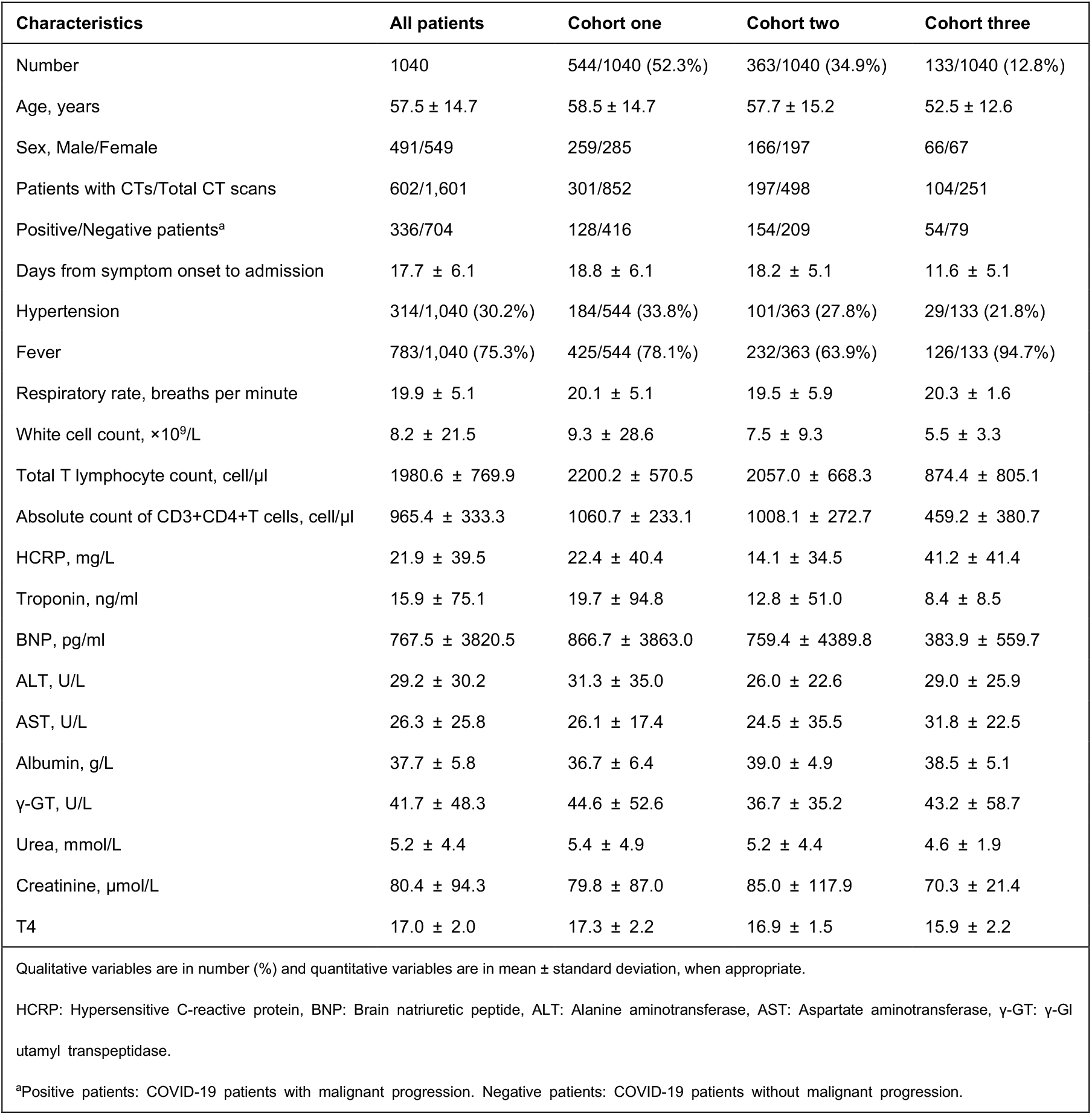
Patient and clinical characteristics.

| **Table 1 Patient and clinical characteristics.** |  |  |  |  |
| --- | --- | --- | --- | --- |
| **Characteristics** | **All patients** | **Cohort one** | **Cohort two** | **Cohort three** |
| Number | 1040 | 544/1040 (52.3%) | 363/1040 (34.9%) | 133/1040 (12.8%) |
| Age, years | 57.5 ± 14.7 | 58.5 ± 14.7 | 57.7 ± 15.2 | 52.5 ± 12.6 |
| Sex, Male/Female | 491/549 | 259/285 | 166/197 | 66/67 |
| Patients with CTs/Total CT scans | 602/1,601 | 301/852 | 197/498 | 104/251 |
| Positive/Negative patients<sup>a</sup> | 336/704 | 128/416 | 154/209 | 54/79 |
| Days from symptom onset to admission | 17.7 ± 6.1 | 18.8 ± 6.1 | 18.2 ± 5.1 | 11.6 ± 5.1 |
| Hypertension | 314/1,040 (30.2%) | 184/544 (33.8%) | 101/363 (27.8%) | 29/133 (21.8%) |
| Fever | 783/1,040 (75.3%) | 425/544 (78.1%) | 232/363 (63.9%) | 126/133 (94.7%) |
| Respiratory rate, breaths per minute | 19.9 ± 5.1 | 20.1 ± 5.1 | 19.5 ± 5.9 | 20.3 ± 1.6 |
| White cell count, ×10<sup>9</sup>/L | 8.2 ± 21.5 | 9.3 ± 28.6 | 7.5 ± 9.3 | 5.5 ± 3.3 |
| Total T lymphocyte count, cell/μl | 1980.6 ± 769.9 | 2200.2 ± 570.5 | 2057.0 ± 668.3 | 874.4 ± 805.1 |
| Absolute count of CD3+CD4+T cells, cell/μl | 965.4 ± 333.3 | 1060.7 ± 233.1 | 1008.1 ± 272.7 | 459.2 ± 380.7 |
| HCRP, mg/L | 21.9 ± 39.5 | 22.4 ± 40.4 | 14.1 ± 34.5 | 41.2 ± 41.4 |
| Troponin, ng/ml | 15.9 ± 75.1 | 19.7 ± 94.8 | 12.8 ± 51.0 | 8.4 ± 8.5 |
| BNP, pg/ml | 767.5 ± 3820.5 | 866.7 ± 3863.0 | 759.4 ± 4389.8 | 383.9 ± 559.7 |
| ALT, U/L | 29.2 ± 30.2 | 31.3 ± 35.0 | 26.0 ± 22.6 | 29.0 ± 25.9 |
| AST, U/L | 26.3 ± 25.8 | 26.1 ± 17.4 | 24.5 ± 35.5 | 31.8 ± 22.5 |
| Albumin, g/L | 37.7 ± 5.8 | 36.7 ± 6.4 | 39.0 ± 4.9 | 38.5 ± 5.1 |
| γ-GT, U/L | 41.7 ± 48.3 | 44.6 ± 52.6 | 36.7 ± 35.2 | 43.2 ± 58.7 |
| Urea, mmol/L | 5.2 ± 4.4 | 5.4 ± 4.9 | 5.2 ± 4.4 | 4.6 ± 1.9 |
| Creatinine, μmol/L | 80.4 ± 94.3 | 79.8 ± 87.0 | 85.0 ± 117.9 | 70.3 ± 21.4 |
| T4 | 17.0 ± 2.0 | 17.3 ± 2.2 | 16.9 ± 1.5 | 15.9 ± 2.2 |
| Qualitative variables are in number (%) and quantitative variables are in mean ± standard deviation, when appropriate. |  |  |  |  |
| HCRP: Hypersensitive C-reactive protein, BNP: Brain natriuretic peptide, ALT: Alanine aminotransferase, AST: Aspartate aminotransferase, γ-GT: γ-Glutamyl transpeptidase. |  |  |  |  |
| <sup>a</sup>Positive patients: COVID-19 patients with malignant progression. Negative patients: COVID-19 patients without malignant progression. |  |  |  |  |

### Performance evaluation and results

Our work advocates the use of a sequence of CT scans, captured at different timings after hospitalization, for accurate malignant progression. Unlike^8,9^, we do not quantify CT scans to avoid information loss, but use a deep learning model to process the raw data directly. In the meantime, an effective integration of CT scans and clinical information underpins our system. The pipeline of our system is given in Fig. 2.

**Fig. 2.**
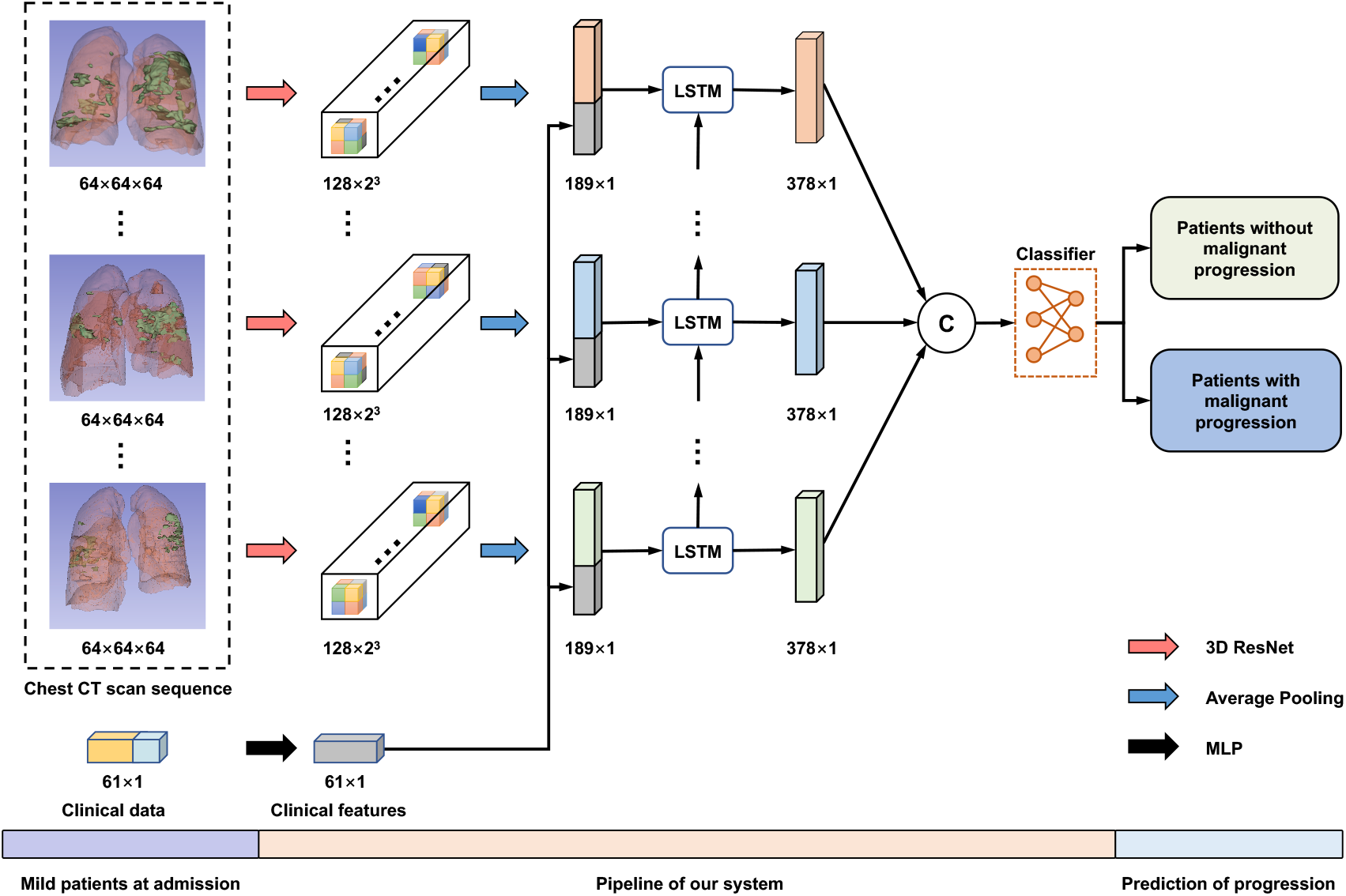
The pipeline of our system about the prediction of COVID-19 malignant progression. First, 3D ResNet and MLP encode chest CT scans and clinical data, respectively. Then, we combine the two features and feed them into an LSTM to model the temporal information. Finally, several fully connected layers are exploited to make the prediction. CT: computed tomography, LSTM: long short-term memory, MLP: multilayer perceptron.

In Table 2, we compare the performance of our model against different methods, including linear discriminant analysis (LDA)^13^, support vector machine (SVM)^14^, and multilayer perceptron (MLP)^12^. In this experiment, we adopt patients with mild symptoms and without malignant progression as the negative reference standard and divide the cohort one into the training cohort (80%) and the validation cohort (20%) randomly. We use five-fold cross-validation to evaluate our model. Our system, which fuses clinical information with the sequential CT scans, achieves a mean AUC of 0.920 (95% CI: [0.861, 0.979]) and outperforms the best traditional machine learning methods (mean AUC of 0.767, 95% CI: [0.725, 0.799]) by a large margin.

**Table 2.** The performance comparison of different methods.

| Methods | AUC | ACC (%) | SEN (%) | SPE (%) | Clinical | Quantitative | CT |
| --- | --- | --- | --- | --- | --- | --- | --- |
|  |  |  |  |  | Data | CT features | scans |
| LDA | 0.675 [0.629, 0.721] | 73.5 [69.8, 77.2] | 20.3 [13.3, 27.3] | 89.9 [87.0, 92.8] | ✓ | × | × |
| LDA | 0.675 [0.629, 0.721] | 67.3 [63.3, 71.2] | 39.1 [30.6, 47.5] | 76.0 [71.9, 80.1] | ✓ | ✓ | × |
| SVM | 0.652 [0.606, 0.699] | 76.3 [72.7, 79.9] | 0.80 [0.00, 2.30] | **99.5 [98.9, 1.00]** | ✓ | × | × |
| SVM | 0.767 [0.725, 0.799] | 76.8 [73.3, 80.4] | 19.5 [12.7, 26.4] | 94.5 [92.3, 96.7] | ✓ | ✓ | × |
| MLP | 0.787 [0.703, 0.872] | 81.6 [78.1, 84.6] | 48.4 [40.0, 57.0] | 91.8 [88.8, 94.1] | ✓ | ✓ | × |
| MLP+3D ResNet | 0.851 [0.775, 0.927] | 85.3 [82.1, 88.0] | 69.5 [61.1, 76.8] | 90.1 [86.9, 92.7] | ✓ | × | ✓ |
| MLP+LSTM | 0.899 [0.836, 0.961] | 86.0 [82.9, 88.7] | 71.9 [63.5, 78.9] | 90.4 [87.2, 92.9] | ✓ | ✓ | × |
| Our System | **0.920 [0.861, 0.979]** | **87.7 [84.7, 90.2]** | **89.1 [82.5, 93.4]** | 87.3 [83.7, 90.1] | ✓ | × | ✓ |

95% confidence intervals are included in brackets.
AUC: area under the receiver operating characteristic curve, ACC: accuracy, SENS: sensitivity, SPEC: specificity, LDA: linear discriminant analysis, SVM: support vector machine, MLP: multilayer perceptron, LSTM: long short-term memory.

According to the results in Table 2, we can draw the following conclusions: 1) CT scans turns out to be more effective than quantitative CT features. For instance, with LSTM, using CT scans obtain a relative improvement of 2.3% over using quantitative CT features in AUC (0.920 vs. 0.899). Without LSTM, the improvement is more distinctive, reaching 8.1% (0.851 vs. 0.787). 2) Modeling the temporal information brings measurable benefits for boosting the performance of our system. This has been evidenced by an improvement of 14.2% (0.899 vs. 0.787) via using LSTM when quantitative CT features are used. Meanwhile, when CT scans are used, the improvement is 8.1% (0.920 vs. 0.851) with a difference of 0.069 in AUC. The corresponding ROCs in Fig. 3a further support our method.

**Fig. 3.**
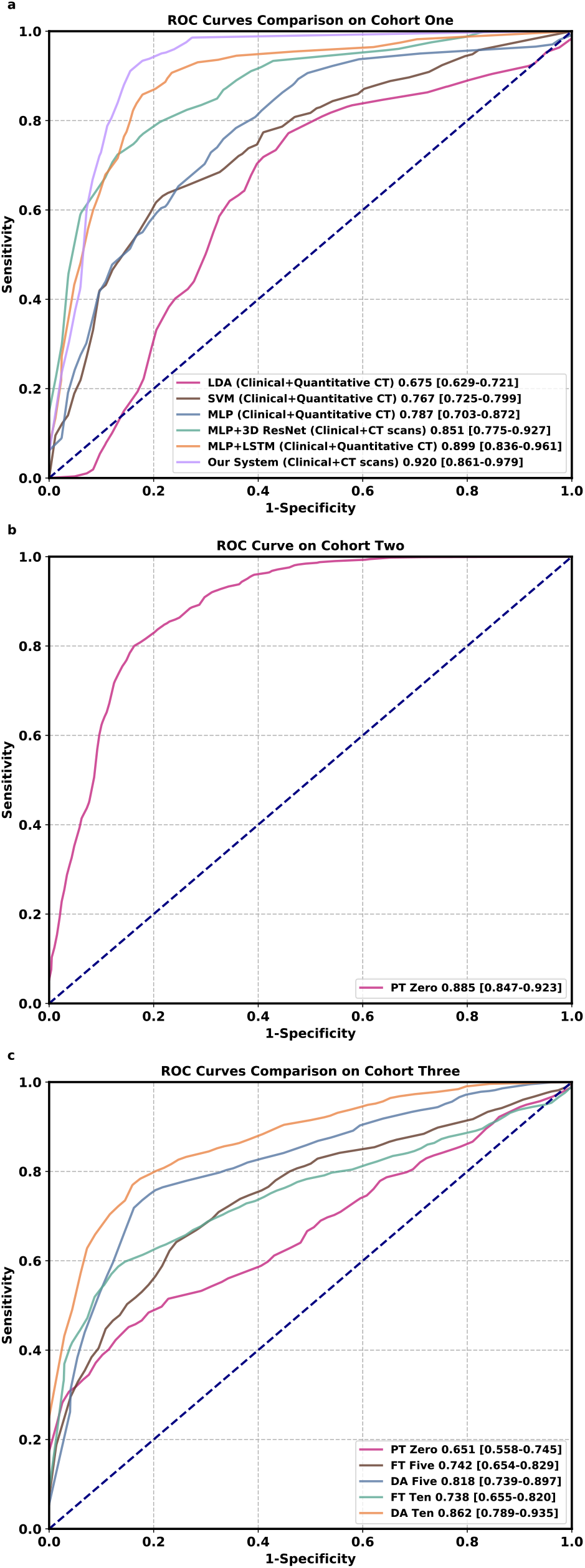
Comparison of ROC curves among different methods on three cohorts. Numbers before brackets are AUCs. Numbers in brackets are confidence intervals. a, ROC curves comparison on cohort one. LDA: linear discriminant analysis, SVM: support vector machine, MLP: multilayer perceptron, LSTM: long short-term memory. b, ROC curve on cohort two. PT: pre-trained. c, ROC curves comparison on cohort three. PT: pre-trained, FT: fine-tuning, DA: domain adaptation. In each class, the same number of samples are used during the domain adaptation process.

### Performance evaluation in the multicenter study

A high-quality labeling process typically requires time-consuming human effort, which is a prominent drawback during the outbreak of COVID-19 where fast analysis is essential. Hence, how to use a small amount of labeled data to improve the generalization power of the system is of great practical significance. Inspired by metric-learning based methods^15^, we propose a domain adaptation method to adapt our model to a new domain with only a few labeled samples available. The details are given in the Methods section.

As Table 3 shows, when directly evaluating the model trained from the cohort one on the cohort two, a satisfactory performance is achieved (AUC: 0.885, 95% CI: [0.847, 0.923]). This is because the two cohorts are from different branches of the same hospital, which means that their data distributions are similar to some degree. However, when directly evaluating the model trained from the cohort one on the cohort three, the performance drops a lot. A mean AUC of 0.651 (95% CI: [0.558, 0.745]) is achieved, because the two cohorts are from different hospitals. When 10 labeled samples in the target domain are used, directly finetuning the model achieves a mean AUC of 0.738 (95% CI: [0.655, 0.820]), still inferior to our system (AUC: 0.862, 95% CI: [0.789, 0.935]). The corresponding ROCs in Fig. 3b,c further support our domain adaptation method.

**Table 3.**
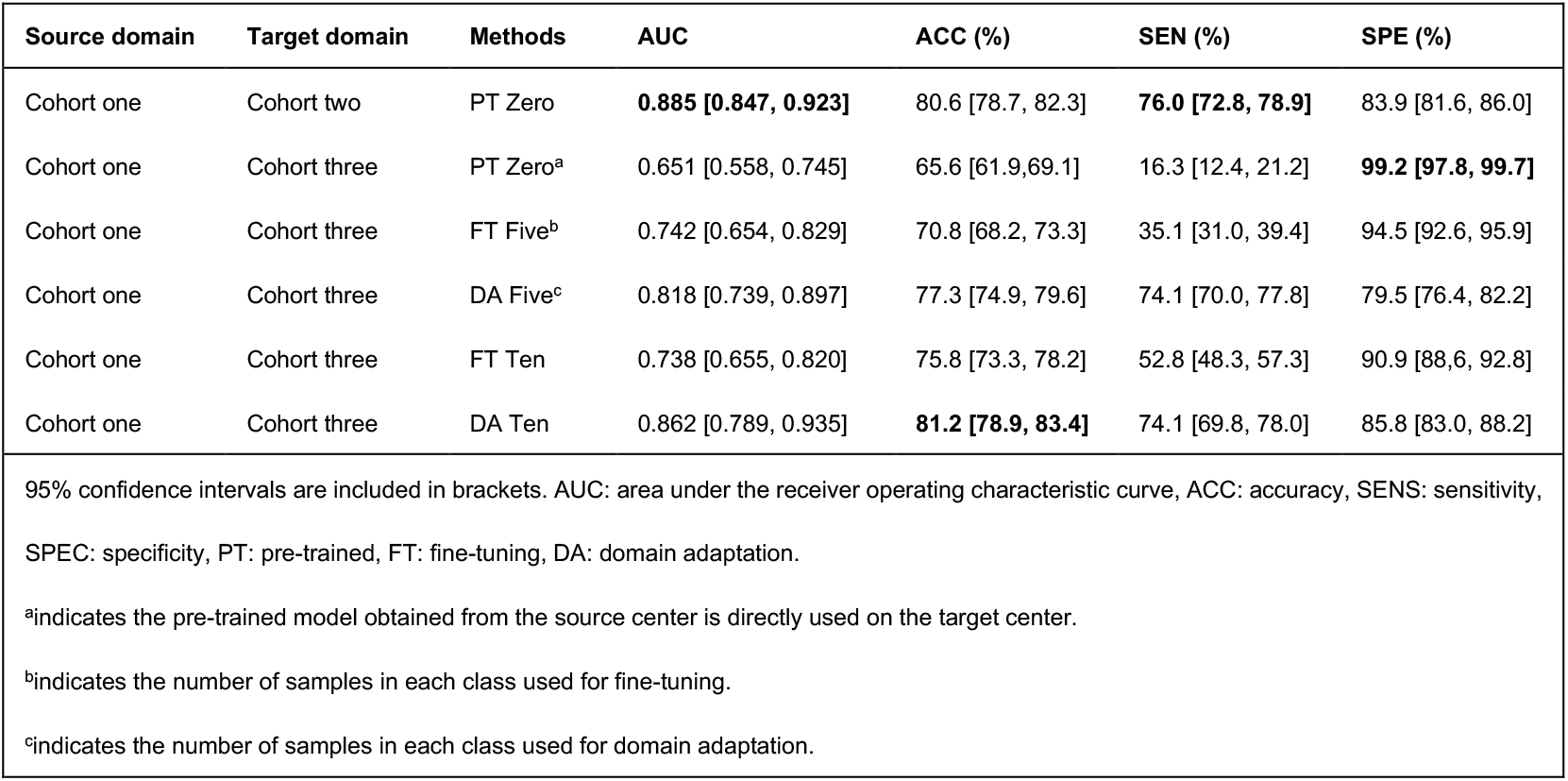
Performance comparison in the multicenter study.

| **Table 3 Performance comparison in the multicenter study.** |  |  |  |  |  |  |
| --- | --- | --- | --- | --- | --- | --- |
| Source domain | Target domain | Methods | AUC | ACC (%) | SEN (%) | SPE (%) |
| Cohort one | Cohort two | PT Zero | **0.885 [0.847, 0.923]** | 80.6 [78.7, 82.3] | **76.0 [72.8, 78.9]** | 83.9 [81.6, 86.0] |
| Cohort one | Cohort three | PT Zero<sup>a</sup> | 0.651 [0.558, 0.745] | 65.6 [61.9, 69.1] | 16.3 [12.4, 21.2] | **99.2 [97.8, 99.7]** |
| Cohort one | Cohort three | FT Five<sup>b</sup> | 0.742 [0.654, 0.829] | 70.8 [68.2, 73.3] | 35.1 [31.0, 39.4] | 94.5 [92.6, 95.9] |
| Cohort one | Cohort three | DA Five<sup>c</sup> | 0.818 [0.739, 0.897] | 77.3 [74.9, 79.6] | 74.1 [70.0, 77.8] | 79.5 [76.4, 82.2] |
| Cohort one | Cohort three | FT Ten | 0.738 [0.655, 0.820] | 75.8 [73.3, 78.2] | 52.8 [48.3, 57.3] | 90.9 [88.6, 92.8] |
| Cohort one | Cohort three | DA Ten | 0.862 [0.789, 0.935] | **81.2 [78.9, 83.4]** | 74.1 [69.8, 78.0] | 85.8 [83.0, 88.2] |
| 95% confidence intervals are included in brackets. AUC: area under the receiver operating characteristic curve, ACC: accuracy, SENS: sensitivity, SPEC: specificity, PT: pre-trained, FT: fine-tuning, DA: domain adaptation. |  |  |  |  |  |  |
| <sup>a</sup>indicates the pre-trained model obtained from the source center is directly used on the target center. |  |  |  |  |  |  |
| <sup>b</sup>indicates the number of samples in each class used for fine-tuning. |  |  |  |  |  |  |
| <sup>c</sup>indicates the number of samples in each class used for domain adaptation. |  |  |  |  |  |  |

### Prognostic factors of clinical data

We further investigate the clinical indicators that contribute to the prediction of the malignant progression by adding a self-attention layer before the first fully connected layer of the MLP. As shown in Supplementary Figure 2, it automatically learns the attention weight corresponding to each clinical indicator, and each attention weight is normalized to (0, 1) by a sigmoid function to measure the importance of the related clinical indicator for the prediction task. The top 20 clinical indicators with the highest attention weights are listed in Supplementary Figure 3. The most important prognostic clinical indicators are myocardial injury (Troponin and Brain natriuretic peptide), followed by hepatic injury (Aspartate aminotransferase, Albumin, and γ-Glutamyl transpeptidase), renal failure (Creatinine), and inflammatory status (Hypersensitive C-reactive protein, White cell count, CD3+CD4+T cells count, fever).

## Discussion

Coronavirus induced pneumonia puts tremendous pressure on public medical systems. Such patients without timely and effective treatment will eventually develop multi-organ failure associated with a high mortality^16,17^. Therefore, early prediction and early aggressive treatment of patients with mild symptoms at a high risk of malignant progression to a severe/critical stage are important ways to reduce mortality.

In this study, we argue that the effective integration of sequential CT scans and clinical data is important for an accurate prediction of malignant progression. Moreover, the rich temporal information contained in the sequence of CT scans, which has not been considered by any studies so far, is critical for this specific task. We have conducted extensive experiments to demonstrate that our system, which effectively fuses the two complementary data, achieves much better performance than using either data as input separately (e.g., 0.851 vs. 0.787 in AUC when using clinical data and quantitative CT features). More importantly, due to the capability of our system in learning temporal information, our system reports a much higher AUC compared with the counterparts that do not consider the temporal information.

Our work is novel because we are among the first attempts to explore ways to fuse clinical data and sequential CT scans to improve the performance of predicting COVID-19 malignant progression in an end to end manner. Experimental results show that both CT scans and clinical data are of paramount importance to this problem. Furthermore, there is little literature concerning the temporal information of CT sequences, however, the temporal cue also contributes significantly to the prediction of malignant progression as it reveals the change of the patient’s health condition.

Traditional machine learning methods heavily rely on domain-specific expertise, as feature patterns to be analyzed are manually designed, which may lead to information loss before feeding them to the classifier. However, our method attempts to automatically learn high-level features from raw data, and jointly optimizes the feature extractor and classifier in an end-to-end manner.

Deep learning-based methods often encounter performance degradation in the multicenter study, mainly due to the large data distribution discrepancy between different cohorts. This is caused by different CT scanners, different slice thickness, different regions, age distribution discrepancy, and systematic errors during the data collection process. Another notable merit of this work is that we employ domain adaptation to improve the robustness of our system in the multicenter study. From comprehensive experimental results, we observe that inferior performance is achieved when the model trained with a single-center is adapted to a completely different data domain by directly fine-tuning. The proposed domain adaptation process enables our system to transfer the prototype representations learned from the source domain to the target domain with a small number of labeled samples, which greatly improves the generalization power in the multicenter evaluation. A well-trained and mature prediction system in one center, which can be quickly deployed in multiple centers, will greatly reduce the significant demand for diagnostic expertise. It effectively optimizes the treatment strategy, thus improving the emergency response capacity of the medical system.

To investigate the interpretability of the CT feature patterns learned by our model, we show the activation maps via using Gradient-weighted Class Activation Mapping (Grad-CAM)^18^. Supplementary Figure 4 shows that the intra-zone and middle-zone of the pulmonary region have the greatest influence on the prediction task, hence are valuable for the predicting of malignant progression.

Our model could effectively identify valuable indicators for predicting the malignant progression of COVID-19 patients with mild symptoms, which assists in clinical assessment and treatment. According to our results, dysfunction or injuries of multiple organs are the most essential predictive indicators for COVID-19 malignant progression, of which myocardial injury is the most important one, followed by liver dysfunction and kidney failure. Coronavirus is currently believed to invade host cells through the angiotensin-converting enzyme 2 (ACE2) pathway to cause COVID-19^19-24^. Since ACE2 is widely distributed in various human tissues, such as type II alveolar cells, myocardial cells, hepatocytes, cholangiocytes, and proximal tubule cells, multiple organ involvement in COVID-19 is not surprising^24-26^. Inflammatory storm caused by coronavirus is another essential predictive indicator for COVID-19 malignant progression. Although the exact mechanisms are unclear yet, patients with COVID-19^27-29^ do show high levels of hypersensitive C-reactive protein and high expression of IL-1B, IFN-γ, IP-10, monocyte chemoattractant protein 1 (MCP-1), etc. These cytokines further activate the T-helper type 1 (Th1) cell response, providing another predictive indicator, CD3+CD4+T cells. The complexity of the clinic and the ambiguity of the pathogenic mechanism significantly increase the difficulty of evaluation and treatment strategy selection for COVID-19 patients.

However, our study still has several limitations. First, samples available for malignant progression prediction are limited. The diverse data in the large-scale dataset will allow deep learning-based methods to gain a more comprehensive understanding of what causes the malignant progression. Second, the data source of our study is limited to three hospital branches of two hospitals in Wuhan. More data needs to be collected from multiple centers, especially from foreign hospitals, to further enhance our model. Third, this study only conducts an interpretable analysis of the relationship between prognostic factors and patients who are easy to deteriorate from the perspective of relevance. Future studies can combine evidence-based medicine to identify the cause and effect of malignant progression.

## Conclusions

In conclusion, our early warning system, built upon the deep learning techniques and the integration of clinical data and sequential CT scans, can accurately predict the malignant progression of COVID-19. Compared with traditional machine learning methods, we demonstrate that our deep learning-based method can learn discriminative feature patterns and improve the prediction performance significantly. Furthermore, the generalization power of our method is improved by domain adaptation in the multicenter study. Our method can identify patients with potentially severe/critical COVID-19 outcomes using an inexpensive, widely available, point-of-care test. Our system can be potentially deployed on the front line to decrease the mortality of COVID-19.

## Methods

### Clinical data acquisition and preprocessing

In Wuhan Pulmonary Hospital, data from 199 patients were collected from January 3, 2020 to February 13, 2020. In Tongji Hospital, data from 2,543 patients were collected from January 13, 2020 to March 16, 2020, of which 544 patients came from the Zhongfa branch, 363 patients came from the Guanggu branch, and 71 patients came from the Main branch. All patients were confirmed by a positive viral nucleic acid test. A subset of 1,040 adult patients belonging to the mild type at admission assessments is selected for further investigation. The inclusion criteria are all of the followings: 1) respiratory rate < 30 breaths per min; 2) resting blood oxygen saturation > 93%; 3) the ratio of arterial oxygen partial pressure to fraction of inspiration oxygen > 300 mm Hg; 4) non-ICU patients without shock, respiratory failure, mechanical ventilation, and failure of other organs. Anyone who fails to fulfill one of the criteria is considered to progress into a severe/critical stage according to the guidelines for the diagnosis and treatment of COVID-19 infection by the Chinese National Health Commission (Version 7)^30^. The medical history, results of physical examination, and laboratory tests were all collected from the HIS system. The time points of symptoms onset and the beginning of severe/critical stage are recorded for further selections of available CT scans. All the patients in the Main branch have mild prognoses, so our research excludes patients from this branch. Furthermore, considering the age imbalance problem, we exclude patients under 18 years old. We finally obtain 61 clinical indicators for each sample.

### CT data acquisition and preprocessing

All the patients underwent serial pulmonary CT exams on dedicated CT scanners (GE, SIEMENS, TOSHIBA, and UNITED IMAGING) in two hospitals with the following parameters: slice thickness 1-3 mm, slice gap 0 mm, 130 kV, 50 mAs. All the CT scans before the severe/critical stage are included to segment the masks of bilateral lungs and pneumonia on an autonomous system (HUAWEI CLOUD Launches AI-Assisted Diagnosis Platform for COVID-19). Each CT scan is downsampled to a 64 × 64 × 64 (width x height x slice) tensor. CT image values are clamped to the range [-1250, 250]. Data augmentations including random horizontal flips and random rotations are used.

As traditional machine learning methods cannot handle raw CT scans directly, we design a set of hand-crafted quantitative features for experimental comparison. These quantitative features include the infection pixels proportion and the average CT value of infection pixels for each CT slice. Finally, a 128-dimensional vector can be obtained for each CT scan. We use zero-padding for the missing CT scan tensor.

### Network architecture and training process

Fig. 2 illustrates the pipeline of our system. As is shown, the input of our model includes the clinical data and a sequence of CT scans obtained at different time points. Specifically, the clinical data is a 61-dimensional vector which is processed by an MLP with identity connections (Supplementary Figure 5). In addition, each CT scan is encoded into a 128-dimensional feature vector by 3D ResNet.

To model the temporal information across the sequence of CT features, we use LSTM for its high capacity in modeling such information^31^ and densely combine the clinical feature and the CT feature via concatenation at each time step. LSTM employed in this study is a single-layer network with an embedding dimension of 189 and a hidden dimension of 378. The output of LSTM, a 378 × 7 tensor, is flattened and then fed into several fully connected layers. Finally, we normalize the output with a softmax layer, which can be interpreted as the probability of the patient’s conversion to the severe/critical stage. The whole model is trained with the cross-entropy loss. The detailed architecture of our model is given in Supplementary Table 1.

### Domain adaptation process

In domain adaptation, we use a metric-based method by using a few labeled samples to bridge the domain gap between the source center (cohort one) and the target center (cohort three). Specifically, our method can be decomposed into two stages: a pre-training stage and a domain adaptation stage. For the pre-training stage, we first train a model on the source center then remove the classifier to get the pre-trained encoder *f_ϕ_*. For the domain adaptation stage, which is the core of our method, we adapt the pre-trained model through a metric-based approach, passing the prototype representations learned from the source center to the target center. The details of this stage are elaborated as follows.

First, we randomly select *N* labeled samples from each class in the target center as the support-set to compute prototypes. At the same time, we randomly choose one sample per class in the target center as the query-set to compute distances to the prototypes in the embedding space. Specifically, in each class let *S* = {(***x****_1_, y_1_*), …, (***x****_N_*,*y_N_*)} denotes a small support-set of *N* labeled samples, where ***x****_i_* is the feature vector of an example and *y_i_* ∊ {0, 1} is the corresponding label. *S_k_* denotes the support-set of examples with class *k* ∊ {0, 1}. We compute the mean vector ***w****_k_* of the embedded support points as prototypes for the two classes:

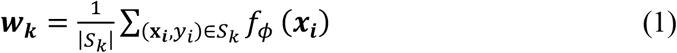

Second, we compute the predicted probability distribution for each sample in the query-set based on a softmax over its cosine similarities (denoted by *sim*(.)) with the prototypes in the embedding space:

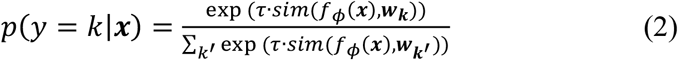

Similar to these studies^32-34^, we also use a learnable parameter *τ* to control the probability distribution sharpness generated by the softmax function during training.

Finally, we train our framework with the classification and similarity losses jointly. Concretely, the classification loss is computed based on *p* and the labels in the query-set:

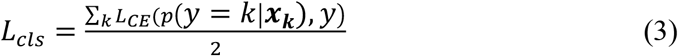

where *L_CE_* is the cross-entropy loss. The similarity loss is adopted to increase the distance between the two class prototypes, which is defined as:

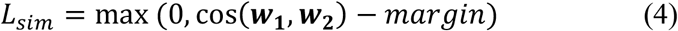

Based on the above two losses, our final loss function is formulated as:

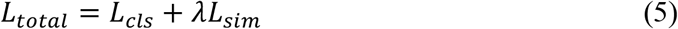

where *λ* is the coefficient to balance the two loss terms.

In the testing stage, all labeled samples in the support-set and the query-set are used to compute prototypes, a test sample will be classified by the similarity between the feature vector of the sample and the prototype of class *k*.

### Implementation details

The proposed network is implemented using Python (Version 3.6 with scipy, scikit-learn, and PyTorch). For single-center experiments on the cohort one, the network is trained by Adam optimizer with an initial learning rate of 0.05, and a batch size of 32 on a single NVIDIA Titan X GPU. The learning rate is decayed by a factor of 10 every 30 epochs. We train our model for 100 epochs. Model weights are initialized with Kaiming method^35^ and bias are initialized as 0. For multicenter experiments on the cohort three, we use the same optimizer settings as single-center experiments, but with a fixed learning rate of 0.01. We finetune 50 epochs for domain adaptation and the batch size is the same as the number of labeled samples with a maximum of 20. The cosine scaling parameter *τ* is initialized to 5 with a fixed learning rate of 0.1. The loss coefficient *λ* is 0.5. The margin of the similarity loss is set to 0.2.

### Performance evaluation criterion

The AUC, accuracy, sensitivity, specificity, and ROC curve are used to evaluate the model performance. The calculation method is shown in Supplementary methods. The 95% bilateral confidence interval is used for all metrics, where the AUC metric uses the Wald-cc interval^36,37^ and the other metrics use the Wilson interval^38^.

## Data Availability

The corresponding author had full access to all data and the final responsibility to submit for publication.

## Data availability

Excel files containing raw data included in the main figures and tables can be found in the Source Data File in the article. All other data are available in the Article and Supplementary Information. All other data including the imaging data can be provided upon reasonable request to the corresponding author.

## Code availability

The code and the pre-trained models are available on GitHub: https://github.com/CongFang/PMP-COVID-19

## Acknowledgments

This work was supported by National Key R&D Program of China (No. 2018YFB1004600), HUST COVID-19 Rapid Response Call (No. 2020kfyXGYJ093, No. 2020kfyXGYJ094), National Natural Science Foundation of China (No.61703049, No. 81401390).

## Author contributions

X.B., C.F., and W.C. conceived the project. C.F. and S.G. developed the machine learning algorithms with the help of S.B., Y.Z., and X.B. Q.C., W.C., L.X., L.Q., and C.H.Z. collected the data. W.C., Q.C., D.T., C.Z.Z., and X.L. analyzed the imaging data. C.F., W.C., Y.Z., S.B., X.X., and Q.C. interpreted the data. C.F., S.B., W.C., X.B., and P.T. wrote the manuscript. W.C., X.B., and P.T. supervised the project. All authors read and approved the final manuscript.

## Additional information

Supplementary information is available for this paper.

